# Community-level Knowledge, Attitudes, and Practices regarding Cardiovascular Diseases and Modifiable Risk Factors in India

**DOI:** 10.1101/2024.02.06.24302426

**Authors:** Kavita Singh, Dimple Kondal, Deepa Mohan, Mareesha Gandral, Sheril Rajan, Viswanathan Mohan, Mohammed K Ali, KM Venkat Narayan, Mark D. Huffman, Dorairaj Prabhakaran, Nikhil Tandon

## Abstract

**Background:** Assessment of knowledge, attitudes, and practices (KAP) regarding cardiovascular diseases (CVD) and cardiovascular risk factors (CVRF) is critical to inform CVD prevention strategies, but limited community-level data exist from developing countries.

**Methods and Results:** We conducted a telephone-based survey nested in the on-going CARRS cohort in Delhi and Chennai, India between January 2021 to February 2021. We randomly selected people with CVRF, but no established CVD and those with existing CVD from the CARRS cohort and assessed their 1) knowledge of CVD symptoms and risk factors, 2) attitude towards non-physician health workers (NPHW) facilitated care and text-messages for healthy lifestyle, and 3) practices regarding monitoring of CVRF. We performed logistic regression analyses to investigate the factors associated with KAP. We interviewed 502 participants (283 with CVRF and 219 with CVD); 45.8% were female, and mean age (SD) was 48.1 (11.2) years. The knowledge of heart attack symptoms, stroke symptoms, and CVRF (>75% correct answers) were: 12.9%, 20.7%, and 17.3%, respectively. Individuals with CVRF had 2.5 times lower knowledge of CVD symptoms compared to those with existing CVD. Acceptability of NPHW-facilitated care and text-messages for healthy lifestyle was 60% and 84%, respectively.

**Conclusions:** The knowledge of CVD symptoms and risk factors is below optimal levels, particularly among individuals at high risk of CVD, unskilled workers, those with lower levels of education and income. Innovative use of NPHW along with mHealth tools could potentially offer solutions to reduce the burden of CVD.

## INTRODUCTION

Cardiovascular disease (CVD) is a leading cause of premature death and disability worldwide. A growing body of evidence indicates that South Asians are at a greater risk of CVD than most other ethnic groups^1^. Several upstream factors (e.g., population ageing, urbanization) and downstream modifiable risk factors (e.g., smoking, unhealthy diet, physical inactivity, obesity, hypertension, diabetes, and dyslipidaemia) together fuel the rising burden of CVD among South Asians. In India, CVD affects more than 70 million people, in their most productive years, with severe socio-economic consequences^2^. Early recognition and response to heart attack and stroke symptoms could reduce delays in hospitalization and mortality. Further, knowledge of, monitoring, and efforts to address modifiable risk factors are necessary prerequisites for CVD prevention strategies to promote behaviour change.

The strong association between behavioural risk factors and CVD onset is widely known, yet substantial potential lies in the prevention, screening, and detection of cardiovascular risk factors (CVRF). Monitoring and addressing CVRFs is critical to prevent or at least mitigate the CVD epidemic. In 2012, the Indian government adopted a national action plan to prevent and control non-communicable chronic diseases^3,4^. This plan defined targets for 2025, including a reduction of smoking (30%), hypertension (25%), and no further increase in diabetes or obesity. India is also playing a pivotal role in terms of reaching global CVD targets as defined in the WHO’s Global Action plan for the prevention and control of NCDs, given the projected future growth and aging of the population^5^. However, it is unclear how the knowledge of CVD symptoms and modifiable risk factors vary among people with existing CVD and those with CVRFs, as community-based data are not available so far for major CVRFs and heart attack and stroke symptoms.

Prior studies assessing knowledge, attitudes, and practices (KAP) in South Asia have mainly focused exclusively on hypertension and diabetes^6–9^. Findings revealed a lack of adequate knowledge about CVRFs with many respondents being unaware of the negative consequences of unhealthy diet, obesity, and physical inactivity. However, data are lacking on the community-level awareness of CVRFs, heart attack and stroke symptoms, and how awareness varies among people with CVRFs and those with existing CVD in India to design efficient CVD prevention strategies^10^. CVD risk reduction requires health systems that efficiently screen and stratify individuals at high risk, provide useful lifestyle modification advice, initiate preventive therapies, and mechanisms to promote lifelong adherence to prescribed drug therapies and lifestyle changes. Strategies such as patient education, task-sharing, and SMS based reminders targeted at improving health behaviours are proven effective in reducing CVD risk and improving adherence to treatment ^11–14^. Despite their benefits, uptake of proven strategies in India has been limited.

With advances in technology, behavioural interventions can potentially be delivered to people at high CVD risk involving trained non-physician health workers (NPHW) using mobile phone (mHealth) applications and text messages, yet limited data exists on the acceptability of these strategies from patients’ perspectives.

Thus, understanding the community-level knowledge of CVD symptoms and CVRFs is an important first step to developing and implementing CVD prevention or care strategies for the high-risk South Asian population. This study examined the **knowledge** of heart attack and stroke symptoms, and modifiable risk factors among people with existing CVD and those with CVRFs in India. Further, we analysed their **attitudes** towards NPHW facilitated care and text message-based reminders for clinic visit or lifestyle advice, and **practices** on monitoring of CVRFs. The study results may assist policy makers to develop and expand ongoing efforts to reduce morbidity, mortality, and associated healthcare costs associated with CVD.

## METHODS

### Study setting and population

Study participants were sampled from the ongoing Centre for Cardiometabolic Risk Reduction in South Asia (CARRS) cohort study^15,16^. Briefly, the CARRS study used a multistage cluster random sampling technique to select households and participants from two megacities in India (Chennai and Delhi) and one in Pakistan (Karachi). When recruited, the CARRS sample populations were representative of each city. The details of the CARRS study methodology have been reported^15^. The baseline data collection for the CARRS cohort occurred in 2010 and thereafter five longitudinal, bi-annual follow-up surveys of the cohort participants have been conducted so far. The current telephone survey to assess knowledge, attitudes and practices (KAP) regarding CVD and modifiable risk factors was conducted alongside the 6^th^ bi-annual survey (September 2020-November 2021) in Chennai and Delhi participants. From the existing cohort, we selected all eligible adults who self-reported CVD (based on a self-reported history of coronary heart disease or stroke). In addition, we selected people at high CVD risk, i.e., those without CVD but presence of at least one CVRF such as hypertension (defined as blood pressure ≥140/ ≥90 mm Hg or self-reported treatment for hypertension)^17^, diabetes (defined as having either HbA1c ≥6.5%, fasting blood glucose ≥126 mg/dL or self-reported use of glucose-lowering medications)^18,19^, or both.

### Study design

A cross-sectional, telephone-based survey was conducted in Delhi and Chennai, India from January 2021 to February 2021 to assess the knowledge, attitudes and practices towards CVD and modifiable risk factors. **Figure 1** shows the selection of study participants from the ongoing cohort and assessment tools.

**Figure 1.**
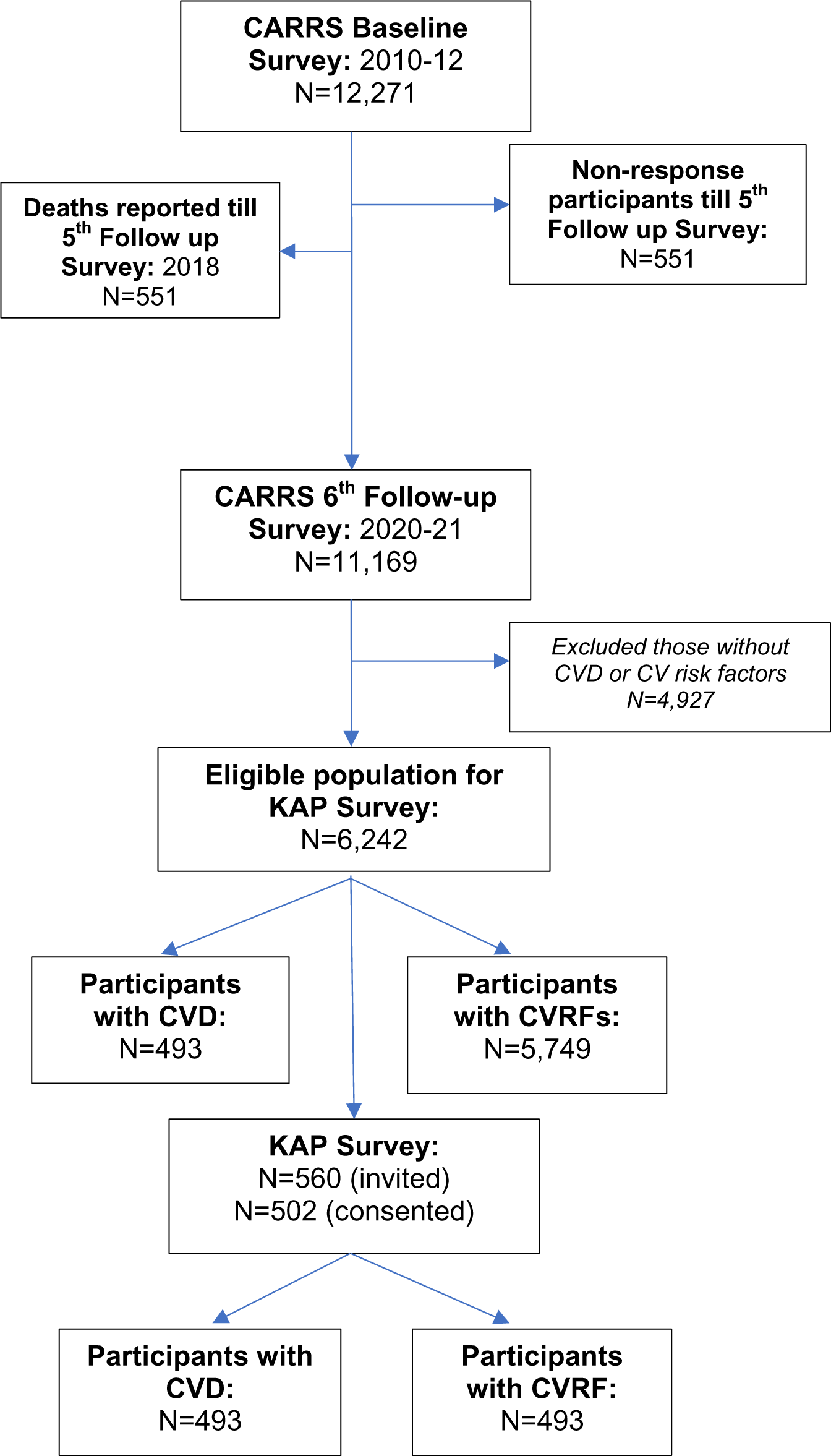
Study flowchart to demonstrate selection of study participants

### Data collection

A structured pre-tested questionnaire was used to collect data on the knowledge of heart attack and stroke symptoms and modifiable CVD risk factors such as smoking, unhealthy diet, physical inactivity, high blood pressure, high cholesterol, high blood glucose, stress, and depression. Components of the study questionnaire were taken from published studies^6,8,20^. Further, we assessed participants’ attitudes or acceptability of NPHWs (nurse, community health workers, or pharmacists) delivered care and willingness to receive text messages to support heart healthy lifestyles. In addition, we collected data on the practices related to CVD management including how frequently participants monitored their blood pressure, blood cholesterol, blood glucose, and body weight.

The questionnaire was initially developed in English and then translated into Hindi. To ensure accuracy of the English to Hindi translation, the questionnaire was back translated from Hindi into English (**Supplement**). The survey questionnaire had additional questions designed to cross-check validity of participants’ responses. In addition, participants’ data on self-reported medical history of hyperlipidaemia, chronic kidney disease, and biomarkers such as body mass index (BMI), blood pressure, blood cholesterol and blood glucose were obtained from the latest available data of CARRS cohort conducted in the fourth follow up (2016-17). We used biomarkers data from the previous follow-up survey because in the current telephone survey conducted between 2020-2021 no blood samples were taken.

Since this study was conducted during the COVID pandemic in India (January 2021-February 2021), we followed COVID-appropriate policies and national guidelines for data collection^21^. The telephone survey was administered by trained field interviewers, and verbal consent was obtained from the study participants. Data were collected using the Commcare application installed in electronic tablets^22^. This study was approved by the institutional ethics committee of the Public Health Foundation of India (TRC-IE No:TRC-IEC-382.1/18) and Madras Diabetes Research Foundation in Chennai, India (Reference no. MDRF/NCT/06-01/2020).

### Data analysis

The demographic and clinical characteristics for the overall study population, and participants with existing CVD and those with CVRFs, are reported as mean (standard deviations, SD) or median (25^th^ and 75^th^ percentiles) for continuous variables and frequencies (percentages) for categorical variables. Knowledge of CVD symptoms and modifiable risk factors are reported as mean (SD) and number (percent) for overall study population and stratified by prior CVD and CVRFs.

Next, we created a composite score to assess the level of knowledge of heart attack and stroke symptoms based on responses to 8 questions. Each correct response was assigned a score of 1 with a total maximal score of 8 points each for the knowledge of heart attack symptoms and stroke symptoms. The total score for heart attack symptoms was classified into three groups: 1 (score: 0-3), 2 (score: 4-5) and 3 (score: 6-8). Likewise, the total score for stroke symptoms was classified into three strata: 1 (score: 0-1), 2 (score: 2-4) and 3 (score: 5-8). Further, knowledge of CVD risk factors was assessed based on responses to 12 questions for a total maximal score of 12 points and was classified into: 1 (score: 0-6), 2 (score: 7-9), 3 (score: 10-12). Finally, we compared the highest vs lowest levels for the knowledge of heart attack symptoms, stroke symptoms, and modifiable risk factors. The lowest level of knowledge scores (i.e., ≤75% correct responses) by people with existing CVD vs those with CVRFs were plotted using bar charts across age-group, sex, and education strata. The chi-square test was performed to investigate the difference in knowledge scores between people with existing CVD vs those with CVRFs across age-group, sex, and education strata.

Bivariate and multivariable regression analyses were performed to assess associations between socio-demographic factors with knowledge (lowest vs highest levels) of heart attack symptoms, stroke symptoms, and modifiable risk factors. Unadjusted and adjusted odds ratios with 95% confidence intervals (CI) are reported. The model was adjusted for age, sex, city, and presence of existing CVD. Next, we analysed the acceptability of NPHW delivered care and willingness to receive text-messages for heart healthy lifestyle by people with CVD and CVRFs. Factors associated with acceptability of NPHW care and text-messages were analysed using bivariable and multivariable logistic regression models. Unadjusted and adjusted odds ratios with 95% CI were calculated, with adjustments for age, sex, city, and presence of existing CVD/CVRFs. Data were available for all covariates and outcomes.

Lastly, the monitoring of CVD risk factors: blood pressure, cholesterol, blood glucose and weight were classified into three groups: 1) measures checked in the past 12 months; 2) measures checked >12 months ago; 3) never checked. Bivariable and multivariable regression analyses were performed to assess the factors associated with monitoring of CVD risk factors in the past 12 months adjusting for age, sex, city, and presence of existing CVD/CVRFs. A two-sided p <0.05 was considered statistically significant without adjustment for multiple testing. All analyses were performed using STATA version 16.0, College Station, Texas, USA.

## RESULTS

### Characteristics of study participants

Between January 2021 – February 2021, 560 participants were invited and 502 participants were consented (89.6% response rate) and completed the telephone survey. The overall mean (SD) age was 48.1 (11.2) years, 45.8% were female, 56.3% (n=283) with CVRF and 43.6% (n=219) with CVD. People with CVRFs were younger (mean age = 45 years vs. 52 years, p<0.001), were more likely to be female (55.1% vs. 33.8%, p<0.001), and less likely to have access to a mobile phone (69.6% vs. 96.3%, p=0.03). Two-thirds of respondents had a high school education, nearly half (47.4%) were unemployed and 54.2% had a monthly household income less than Rs. 10,000. Two-thirds of respondents had internet access (62.3%) on mobile phones. Less than one-quarter had a family history of coronary heart disease (15.5%), with no difference among people with CVRFs and existing CVD (13.4% vs. 18.3%, p=0.14). Overall, half of the study participants had a self-reported history of hypertension (51.1%) with lower rates for diabetes (43.4%) and hyperlipidaemia (38.6%). Among people with CVD, 192 (87.7%) reported coronary heart disease and 38 (17.4%) had a stroke, and 11 (5.0%) had both heart disease and stroke. Further, among people with CVD, 64.8% had hypertension, and 49.3% had diabetes. In contrast, people with CVRFs, 40.6% had hypertension and 38.9% had diabetes (**Table 1**).

**Table 1.** Characteristics of study participants

|  | **OVERALL**<br>**N=502 (%)** | **People with**<br>**CVRF**<br>**N=283 (%)** | **People with**<br>**existing CVD**<br>**N=219 (%)** | **P-value** |
| --- | --- | --- | --- | --- |
| **Age**, in years, mean (SD) | 48.1 (11.2) | 45.0 (10.8) | 52.0 (10.5) | <0.001 |
| **Sex** |  |  |  | <0.001 |
| Male | 272 (54.2%) | 127 (44.9%) | 145 (66.2%) |  |
| Female | 230 (45.8%) | 156 (55.1%) | 74 (33.8%) |  |
| **Marital status** |  |  |  | 0.240 |
| Married | 472 (94.0%) | 263 (92.9%) | 209 (95.4%) |  |
| Non-Married | 30 (6.0%) | 20 (7.1%) | 10 (4.6%) |  |
| **Education** |  |  |  | 0.110 |
| Up to primary schooling | 61 (12.2%) | 28 (9.9%) | 33 (15.1%) |  |
| High school to Secondary | 322 (64.1%) | 181 (64.0%) | 141 (64.4%) |  |
| Graduation and above | 119 (23.7%) | 74 (26.1%) | 45 (20.5%) |  |
| **Occupation** |  |  |  | 0.210 |
| Unemployed | 238 (47.4%) | 134 (47.3%) | 104 (47.5%) |  |
| Semiskilled/Unskilled | 88 (17.5%) | 43 (15.2%) | 45 (20.5%) |  |
| Trained/skilled | 176 (35.1%) | 106 (37.5%) | 70 (32.0%) |  |
| **Household Income (monthly), Indian rupees (US\$)** | | | | 0.530 |
| <10000 (<125) | 272 (54.2%) | 160 (56.5%) | 112 (51.1%) |  |
| 10000-20000 (125-250) | 102 (20.3%) | 56 (19.8%) | 46 (21.0%) |  |
| >20000 (>250) | 126 (25.1%) | 67 (23.7%) | 59 (26.9%) |  |
| Missing | 2 (0.4%) | 0 (0.0%) | 2 (0.9%) |  |
| **Religion** |  |  |  | <0.001 |
| Hindu | 425 (84.7%) | 252 (89.0%) | 173 (79.0%) |  |
| Muslim | 32 (6.4%) | 12 (4.2%) | 20 (9.1%) |  |
| Sikh | 11 (2.2%) | 1 (0.4%) | 10 (4.6%) |  |
| Christian | 32 (6.4%) | 18 (6.4%) | 14 (6.4%) |  |
| Jain | 2 (0.4%) | 0 (0.0%) | 2 (0.9%) |  |
| **City** |  |  |  | <0.001 |
| Chennai | 284 (56.6%) | 183 (64.7%) | 101 (46.1%) |  |
| Delhi | 218 (43.4%) | 100 (35.3%) | 118 (53.9%) |  |
| **Having access to mobile phone** | **408 (81.3%)** | **197 (69.6%)** | **211 (96.3%)** | 0.027 |
| Basic Phone | 156 (38.2%) | 66 (33.5%) | 90 (42.7%) |  |
| Smart Phone | 252 (61.8%) | 131 (66.5%) | 121 (57.3%) |  |
| Having internet access in mobile phone | 254 (62.3%) | 131 (66.5%) | 123 (58.3%) | 0.160 |
| Family history of coronary heart disease | 78 (15.5%) | 38 (13.4%) | 40 (18.3%) | 0.140 |
| Self-reported previous heart disease | 192 (38.2%) | 0 | 192 (87.7%) | <0.001 |
| If yes, taking medications for heart disease (n=192) | 174 (90.6%) | - | 174 (90.6%) |  |
| Self-reported previous stroke | 38 (7.6%) | 0 | 38 (17.4%) | <0.001 |
| If yes, taking medications for stroke (n=38) | 15 (39%) | - | 15 (39%) |  |
| Hypertension | 257 (51.2%) | 115 (40.6%) | 142 (64.8%) | <0.001 |
| If yes, taking medications for hypertension (n=257) | 235 (91.4%) | 102 (88.7%) | 133 (93.7%) | 0.160 |
| Diabetes mellitus | 218 (43.4%) | 110 (38.9%) | 108 (49.3%) | 0.019 |
| If yes, taking medications for diabetes (n=218) | 210 (96.3%) | 106 (96.4%) | 104 (96.3%) | 0.980 |
| Hyperlipidemia | 194 (38.6%) | 67 (23.7%) | 127 (58.0%) | <0.001 |
| If yes, taking medications for hyperlipidemia (n=194) | 156 (80.4%) | 45 (67.2%) | 111 (87.4%) | <0.001 |
| Chronic Kidney Disease (CKD) | 15 (3.0%) | 3 (1.1%) | 12 (5.5%) | 0.004 |
| If yes, taking medications for CKD (n=15) | 10 (67%) | 3 (100%) | 7 (58%) | 0.170 |
| Body mass index*, Kg/m<sup>2</sup> mean (SD) (N=423) | 26.5 (4.6) | 26.3 (4.6) | 26.7 (4.6) | 0.440 |
| Systolic blood pressure*, mmHg (N=469) | 130.4 (18.5) | 130.4 (18.4) | 130.5 (18.7) | 0.960 |
| Diastolic blood pressure*, mmHg (N=469) | 81.8 (11.4) | 81.7 (11.2) | 82.0 (11.7) | 0.76 |
| Total cholesterol*, mean (SD) (N=460) | 176.9 (42.2) | 183.3 (37.8) | 168.7 (46.1) | <0.001 |
| Fasting blood glucose*, mean (SD) (N=461) | 127.6 (54.8) | 124.8 (53.8) | 131.4 (56.1) | 0.200 |

\*BMI, blood pressure, cholesterol and blood glucose data obtained from the fourth follow-up of CARRS cohort conducted in 2016-17.
Marital status, Religion and Family history of coronary heart disease data obtained from CARRS baseline survey conducted in 2010-11
CVRF=Cardiovascular Risk Factors, CVD=Cardiovascular Disease, INR=Indian rupees; SD=Standard Deviation
P-value reports the difference between people with CVRF vs people with CVD.

### Knowledge of heart attack and stroke symptoms and modifiable CVD risk factors

The overall mean (SD) knowledge score for heart attack symptoms was 3.9 (2.3), which was significantly lower among people with CVRFs than those with CVD (3.2 vs. 4.8, p<0.001). Further, knowledge of heart attack symptoms (defined as >75% correct responses) was 12.9%, which was significantly lower among people with CVRFs than those with CVD (8.5% vs. 18.7%, p<0.01). The mean (SD) knowledge score for stroke symptoms was lower at 2.8 (2.5), with no difference between people with CVRFs and CVD (p=0.70). Further, knowledge of stroke symptoms (>75% correct response) was 20.7%, with no difference between those with risk factors or existing CVD (p = 0.60). The mean (SD) knowledge of modifiable risk factors was 7 (2.6) with no significant difference between people with CVRFs and CVD (p=0.04). Overall, the knowledge of modifiable risk factors (>75% correct responses) was 17.3%, with similar proportions among people with CVRFs and CVD (p=0.47). In terms of knowledge of single modifiable risk factors, most participants recognized high cholesterol (88.6%), stress (82.9%), and hypertension (81.1%) as the factors that increases the risk of having a heart attack, followed by unhealthy diet (64.5%), diabetes (63.1%), smoking (60.6%) and obesity (59.6%). Less than one-quarter of study participants reported that daily exercise (22.7%) and sleeping too much (15.1%) increased the risk of a heart attack, which was similar between people with CVRFs and CVD except for an increased awareness among people with CVD vs CVRFs for smoking (66.2% vs. 56.2%, p=0.02), alcohol use in excess (63.9% vs. 55.5%, p=0.05), and diabetes (69.4% vs. 58.3%, p=0.01) (**Table 2**).

**Table 2.**
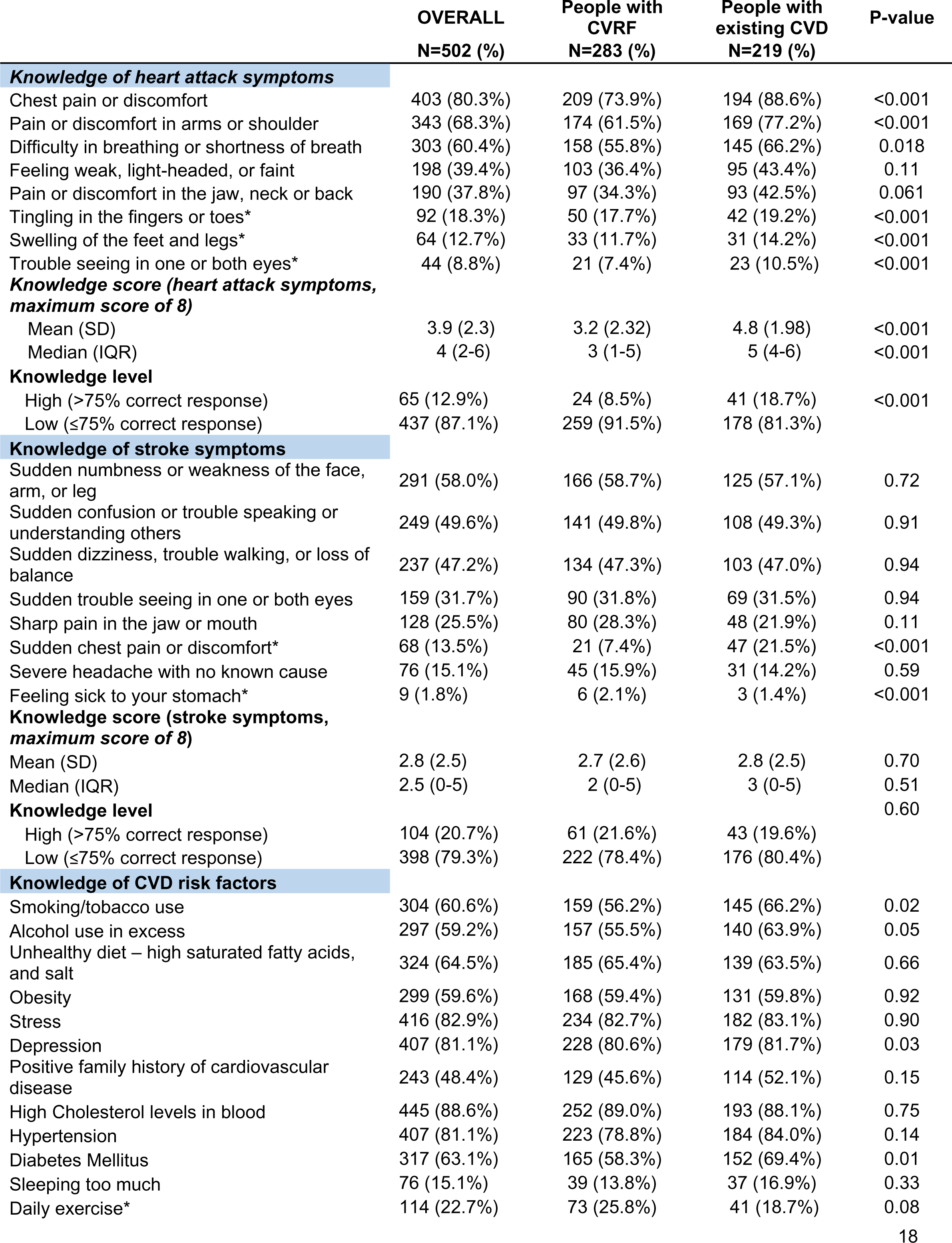
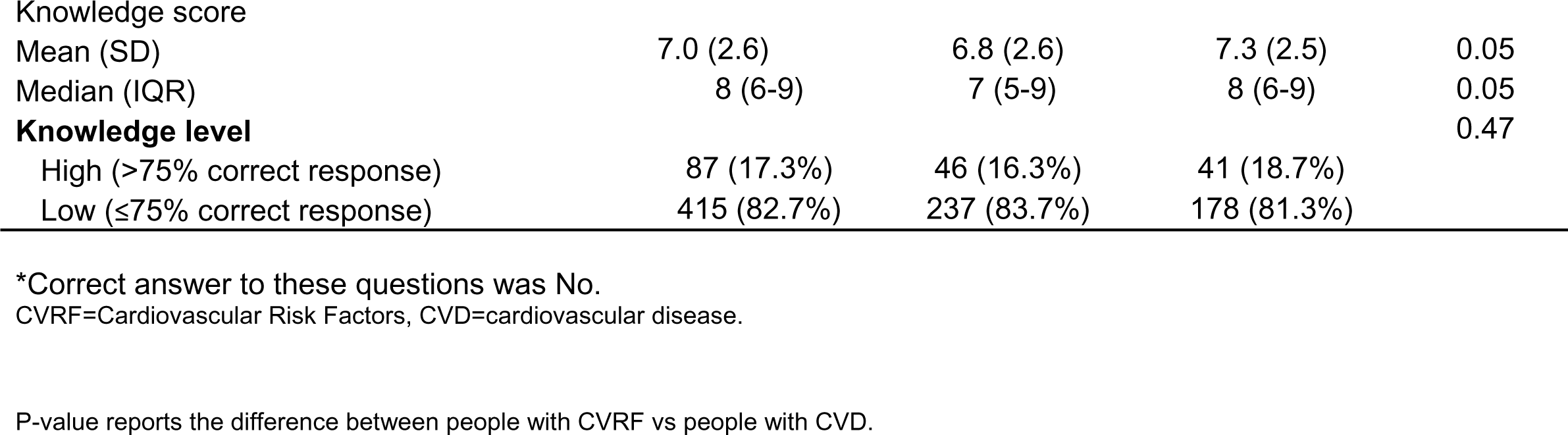
Knowledge of heart attack symptoms, stroke symptoms and cardiovascular risk factors

The bivariate analysis comparing knowledge of heart attack symptoms, stroke symptoms and modifiable CVD risk factors between people with CVRF and CVD across age, sex, and education groups are shown in **e-Figures 1a-1c**. Overall, younger age (<35 years) and elderly (>55 years) versus middle aged participants (35-55 years)), females vs. males, were more likely to have low knowledge of heart attack symptoms, stroke symptoms, and modifiable CVD risk factors (p<0.001).

In the adjusted multivariable regression model (**Table 3**), lower knowledge of heart attack symptoms were predominantly found in people with CVRF vs people with CVD (OR=2.57, 95% CI: 1.43, 4.62), and those with lower levels of education and unemployed, but the latter did not reach statistical significance.

**Table 3.** Factors associated with the knowledge of heart attack and stroke symptoms, and risk factors.

|  | Knowledge score |  | Unadjusted | Adjusted* |
| --- | --- | --- | --- | --- |
|  | High<br>N (%) | Low<br>N (%) | Odds ratio<br>(95% CI) | Odds ratio<br>(95% CI) |
| **Knowledge of heart attack symptoms** |  |  |  |  |
| People with CVRF | 24 (8.5%) | 259 (91.5%) | 2.49 (1.45,4.26) | 2.57 (1.43,4.62) |
| Female | 24 (10.4%) | 206 (89.6%) | 1.52 (0.89,2.61) | 1.28 (0.74,2.24) |
| Age | 65 (13.0%) | 437 (87.0%) | 1.01 (0.79, 1.28) | 1.02 (0.99,1.05) |
| Marital Status (Married) | 61 (12.9%) | 411 (87.1%) | 1.04 (0.35,3.07) | 1.29 (0.42,3.96) |
| Education |  |  |  |  |
| *Graduation and above* | 17 (14.3%) | 102 (85.7%) | ref | ref |
| *Up to primary schooling* | 6 (9.8%) | 55 (90.2%) | 1.53 (0.57,4.1) | 1.47 (0.52,4.14) |
| *High school to Secondary school* | 42 (13%) | 280 (87.0%) | 1.11 (0.61,2.04) | 1.04 (0.54,2.02) |
| Occupation |  |  |  |  |
| *Trained/skilled* | 26 (14.8%) | 150 (85.2%) | ref | ref |
| *Unemployed* | 25 (10.5%) | 213 (89.5%) | 1.48 (0.82,2.66) | 1.36 (0.60,3.06) |
| *Semiskilled/Unskilled* | 14 (15.9%) | 74 (84.1%) | 0.92 (0.45,1.86) | 0.96 (0.45,2.02) |
| Monthly household income (in Indian rupees) |  |  |  |  |
| >20,000 | 15 (11.9%) | 111 (88.1%) | ref | ref |
| <10,000 | 32 (11.8%) | 240 (88.2%) | 1.01 (0.53,1.95) | 0.78 (0.37,1.67) |
| 10,000-20,000 | 16 (15.7%) | 86 (84.3%) | 0.73 (0.34,1.55) | 0.62 (0.28,1.37) |
| Family history of CHD (No) | 52 (12.3%) | 372 (87.7%) | 1.43 (0.74,2.77) | 1.26 (0.64,2.50) |
| **Knowledge of symptoms of stroke** |  |  |  |  |
| People with CVRF | 61 (21.6%) | 222 (78.4%) | 0.89 (0.57,1.38) | 0.65 (0.38,1.09) |
| Female | 50 (21.7%) | 180 (78.3%) | 0.89 (0.58,1.37) | 0.86 (0.53,1.40) |
| Age | 104 (20.7%) | 398 (79.2%) | 1.11 (0.91, 1.36) | 1.01 (0.99,1.04) |
| Marital Status (Married) | 99 (21.0%) | 373 (79.0%) | 0.75 (0.28,2.02) | 0.58 (0.2,1.66) |
| Education |  |  |  |  |
| *Graduation and above* | 45 (37.8%) | 74 (62.2%) | ref | ref |
| *Up to primary schooling* | 8 (13.1%) | 53 (86.9%) | 4.03 (1.76,9.24) | 2.05 (0.83,5.05) |
| *High school to Secondary school* | 51 (15.8%) | 271 (84.2%) | 3.23 (2.01,5.20) | 1.53 (0.90,2.61) |
| Occupation |  |  |  |  |
| *Trained/skilled* | 51 (29.0%) | 125 (71.0%) | ref | ref |
| *Unemployed* | 46 (19.3%) | 192 (80.7%) | 1.70 (1.08,2.69) | 1.61 (0.79,3.29) |
| *Semiskilled/Unskilled* | 7 (8.0%) | 81 (92.0%) | 4.72 (2.04,10.91) | 3.43 (1.39,8.43) |
| Monthly household income (in Indian rupees) |  |  |  |  |
| >20,000 | 51 (40.5%) | 75 (59.5%) | ref | ref |
| <10,000 | 29 (10.7%) | 243 (89.3%) | 5.70 (3.37,9.62) | 2.81 (1.54,5.13) |
| 10,000-20,000 | 24 (23.5%) | 78 (76.5%) | 2.21 (1.24,3.95) | 1.56 (0.83,2.91) |
| Family history of CHD (No) | 79 (18.6%) | 345 (81.4%) | 2.06 (1.21,3.52) | 1.46 (0.81,2.65) |
| **Knowledge of modifiable CVD risk factors** |  |  |  |  |
| People with CVRF | 46 (16.3%) | 237 (83.7%) | 1.19 (0.75,1.89) | 0.99 (0.58,1.7) |
| Female | 39 (17.0%) | 191 (83.0%) | 1.05 (0.66,1.67) | 1.00 (0.61,1.65) |
| Age | 87 (17.3%) | 415 (82.7%) | 1.06 (0.85, 1.31) | 1.02 (0.99,1.04) |
| Marital Status (Married) | 78 (16.5%) | 394 (83.5%) | 2.16 (0.96,4.90) | 2.16 (0.88,5.28) |
| Education |  |  |  |  |
| *Graduation and above* | 37 (31.1%) | 82 (68.9%) | ref | ref |
| *Up to primary schooling* | 10 (16.4%) | 51 (83.6%) | 2.30 (1.05,5.03) | 1.26 (0.54,2.94) |
| *High school to Secondary school* | 40 (12.4%) | 282 (87.6%) | 3.18 (1.91,5.30) | 1.76 (1.01,3.07) |
| Occupation |  |  |  |  |
| *Trained/skilled* | 38 (21.6%) | 138 (78.4%) | ref | ref |
| *Unemployed* | 39 (16.4%) | 199 (83.6%) | 1.41 (0.86,2.31) | 1.02 (0.49,2.10) |
| *Semiskilled/Unskilled* | 10 (11.4%) | 78 (88.6%) | 2.15 (1.01,4.55) | 1.45 (0.65,3.23) |
| Monthly household income (in Indian rupees) |  |  |  |  |
| *>20,000* | 36 (28.6%) | 90 (71.4%) | ref | ref |
| *<10,000* | 27 (9.9%) | 245 (90.1%) | 3.63 (2.08,6.32) | 1.84 (0.98,3.45) |
| *10 000-20,000* | 24 (23.5%) | 78 (76.5%) | 1.30 (0.71,2.37) | 0.91 (0.48,1.72) |
| Family history of CHD (No) | 69 (16.3%) | 355 (83.7%) | 1.54 (0.86,2.77) | 1.09 (0.58,2.04) |

The model is adjusted for age, sex, city, and presence of existing CVD/CVRFs CVRF=Cardiovascular Risk Factors, CHD=coronary heart disease

The lower knowledge of stroke symptoms was prevalent among semiskilled/unskilled workers vs trained/skilled workers (OR=3.43, 95% CI: 1.39, 8.43). Further, there were generally no significant differences in terms of the knowledge of modifiable risk factors by age, sex, income categories, except lower education was associated with low knowledge of modifiable CVD risk factors (high school vs. graduates; OR=1.76, 95% CI: 1.01-3.07).

### Attitude towards involving non-physician health workers and use of text-message in CVD care

More than two-thirds of respondents (72%) faced difficulty in understanding written medical information received from the hospital, with significantly higher problems reported by people with CVD than those with CVRFs (78.5% vs. 64.6%, p=0.002). Further, more than half of respondents (60%) were willing to receive counselling on healthy diet, exercise, prescribed medicines, and clinic visit reminders offered through a trained non-physician health worker (NPHW) such as a nurse or community health worker, with no significant differences between people with CVD and CVRFs. Nearly half of participants were willing to have their blood pressure, blood cholesterol, and blood glucose measured by a trained NPHW, and one-third preferred receiving reminders for laboratory appointments, with significantly higher acceptance among people with CVD than those with CVRFs (p <0.001). One-third of participants preferred receiving advice on smoking cessation from NPHWs, with no significant difference between groups (p=0.08). Lastly, most respondents (84%) were willing to receive text-messages about heart healthy lifestyle, with significantly higher acceptance among people with CVRFs vs with CVD (88.3% vs. 80.5%, p=0.03) (**Table 4**).

**Table 4.** Attitudes towards the non-physician health workers and text messages for cardiovascular disease received from the hospital or medical notes

|  | Overall<br>N=502 (%) | People with<br>CVRF<br>(N=283) (%) | People with<br>existing CVD<br>(N=219) (%) | P-value |
| --- | --- | --- | --- | --- |
| Difficulty in understanding written information received from the hospital | 300 (71.9%) | 128 (64.6%) | 172 (78.5%) | 0.002 |
| Need assistance to read physician prescription or medical notes | 304 (72.9%) | 113 (57.1%) | 191 (87.2%) | <0.001 |
| **Acceptability of NPHW delivered care** |  |  |  |  |
| Offer advice on healthy diet | 248 (59.9%) | 122 (61.9%) | 126 (58.1%) | 0.42 |
| Offer advice on exercise | 246 (59.3%) | 126 (63.6%) | 120 (55.3%) | 0.08 |
| Offer advice on smoking cessation | 142 (34.2%) | 76 (38.4%) | 66 (30.4%) | 0.08 |
| Measure blood pressure | 212 (51.1%) | 76 (38.4%) | 136 (62.7%) | <0.001 |
| Measure blood glucose | 198 (47.7%) | 68 (34.3%) | 130 (59.9%) | <0.001 |
| Measure blood cholesterol | 192 (46.3%) | 63 (31.8%) | 129 (59.4%) | <0.001 |
| Helping patients managing their prescribed medicine | 241 (58.1%) | 127 (64.1%) | 114 (52.5%) | 0.01 |
| Reminder for clinic visits | 250 (60.2%) | 142 (71.7%) | 108 (49.8%) | <0.001 |
| Reminder for lab appointments | 150 (36.1%) | 57 (28.8%) | 93 (42.9%) | 0.01 |
| **Willing to receive text messages on healthy lifestyle** | 344 (84.3%) | 174 (88.3%) | 170 (80.57%) | 0.03 |
| **Reasons for not opting text messages (N=64)** |  |  |  |  |
| Too expensive | 4 (6.2%) | 0 (0%) | 4 (9.9%) | 0.26 |
| Not worried about my health | 4 (6.3%) | 2 (3.2%) | 2 (4.9%) |  |

NPHW=non-physician health worker
The model is adjusted for age, sex, city, and presence of existing CVD/CVRFs
CVRF=Cardiovascular Risk Factors, CVD=cardiovascular disease

Participants from Delhi vs Chennai reported significantly greater acceptability towards NPHW delivered care (OR=9.3, 95%CI: 4.54,19.03) and willingness to receive text-messages for healthy lifestyle (OR=6.79, 95%CI: 2.84,16.21). People with CVRFs vs those with CVD had two times greater acceptability to receive text messages for healthy lifestyle (OR=2.25, 95%CI: 1.19, 4.26) (**Table 5**).

**Table 5.** Factors associated with acceptability of non-physician health worker and text messages for cardiovascular disease management.

|  | Acceptability of NPHW facilitated care |  | Unadjusted | Adjusted* |
| --- | --- | --- | --- | --- |
|  | No<br>N (%) | Yes<br>N (%) | Odds ratio (95% CI) | Odds ratio (95% CI) |
| **Acceptability of NPHW for any services** |  |  |  |  |
| People with CVRF | 47 (23.7%) | 151 (76.3%) | 0.99 (0.63,1.55) | 0.92 (0.55,1.53) |
| City (Delhi) | 24 (24.5%) | 191 (60.3%) | 4.67 (2.80,7.80) | 9.30 (4.54,19.03) |
| Female | 45 (24.7%) | 137 (75.3%) | 0.90 (0.57,1.41) | 0.89 (0.54,1.47) |
| Age (in years) | 98 (23.6%) | 317 (76.4%) | 0.97 (0.95,0.99) | 0.97 (0.95,0.99) |
| Marital Status (Married) | 93 (23.9%) | 297 (76.1%) | 0.80 (0.29,2.19) | 0.76 (0.25,2.32) |
| Education |  |  |  |  |
| *Graduation and above* | 19 (18.1%) | 86 (81.9%) | ref | ref |
| *Up to primary schooling* | 13 (26.0%) | 37 (74.0%) | 0.63 (0.28,1.40) | 1.79 (0.69,4.65) |
| *High school to Secondary school* | 66 (25.4%) | 194 (74.6%) | 0.65 (0.37,1.15) | 1.72 (0.86,3.45) |
| Occupation |  |  |  |  |
| *Trained/skilled* | 26 (17.9%) | 119 (82.1%) | ref | ref |
| *Unemployed* | 55 (27.4%) | 146 (72.6%) | 0.58 (0.34, 0.98) | 0.94 (0.433, 2.05) |
| *Semiskilled/Unskilled* | 17 (24.6%) | 52 (75.4%) | 0.67 (0.33,1.34) | 1.01 (0.48,2.13) |
| Monthly household income (in Indian rupees) |  |  |  |  |
| >20 000 | 25 (21.5%) | 91 (78.5%) | ref | ref |
| <10000 | 98 (23.7%) | 315 (76.3%) | 0.85 (0.50, 1.47) | 3.22 (1.50, 6.93) |
| 10000-20000 | 22 (25.0%) | 66 (75.0%) | 0.82 (0.43,1.59) | 1.83 (0.83,4.04) |
| Family history of CHD (No) | 87 (25.1%) | 259 (74.9%) | 0.56 (0.28,1.12) | 0.79 (0.38,1.64) |
| **Willing to receive text messages about heart healthy living** |  |  |  |  |
| People with CVRF | 23 (11.6%) | 174 (88.4%) | 1.82 (1.05,3.17) | 2.25 (1.19,4.26) |
| City (Delhi) | 9 (14.1%) | 203 (59.0%) | 8.8 (4.21,18.38) | 6.79 (2.84,16.21) |
| Female | 32(18.4%) | 142 (81.6%) | 0.70 (0.41,1.20) | 0.53 (0.29,0.98) |
| Age (in years) | 64 (15.7%) | 344 (84.3%) | 0.96 (0.94,0.98) | 0.96 (0.93,0.98) |
| Marital Status (Married) | 61 (15.9%) | 323 (84.1%) | 0.76 (0.22,2.61) | 0.67 (0.16,2.81) |
| Education |  |  |  |  |
| *Graduation and above* | 4 (3.8%) | 101 (96.2%) | ref | ref |
| *Up to primary schooling* | 15 (31.3%) | 33 (68.7%) | 0.09 (0.03,0.28) | 0.32 (0.09,1.16) |
| *High school to Secondary school* | 45 (17.6%) | 210 (82.0%) | 0.18 (0.06,0.53) | 0.57 (0.18,1.8) |
| Occupation |  |  |  |  |
| *Trained/skilled* | 9 (6.2%) | 137 (93.8%) | ref | ref |
| *Unemployed* | 39 (20.2%) | 154 (79.8%) | 0.26 (0.12,0.55) | 0.61 (0.22,1.71) |
| *Semiskilled/Unskilled* | 16 (23.2%) | 53 (76.8%) | 0.22 (0.09,0.52) | 0.38 (0.15,0.97) |
| Monthly household income (in Indian rupees) |  |  |  |  |
| >20,000 | 5 (4.4%) | 109 (95.6%) | ref | ref |
| <10,000 | 47 (22.9%) | 158 (77.1%) | 0.15 (0.06,0.40) | 0.41 (0.14,1.24) |
| 10,000-20,000 | 11 (12.6%) | 76 (87.4%) | 0.32 (0.11,0.95) | 0.61 (0.19,2.01) |
| Family history of CHD (No) | 61 (18.0%) | 278 (82.0%) | 0.21 (0.06,0.68) | 0.25 (0.07,0.88) |

\*Each covariate is adjusted with city, age and sex
CVRF=Cardiovascular Risk Factors, CHD=coronary heart disease

### Practices related to the monitoring of CVD risk factors

Overall, less than one-third of the participants had their blood pressure, blood glucose, body weight, and blood cholesterol checked >12 months ago and Less than 10% of the study participants had never checked any of these measures (**eTable 1**). The adjusted logistic regression model showed that people with lower levels of education (OR=2.84, 95%CI: 1.39, 5.78) and income (OR=2.02, 95%CI: 1.19, 3.41) had poor monitoring of cardiovascular risk factors (**eTable 2**).

## DISCUSSION

To the best of our knowledge, this is the first community-level data documenting variation in knowledge scores for heart attack and stroke symptoms and modifiable risk factors among adults with CVD compared to those at high CVD risk in two megacities in India. In this study, knowledge of heart attack symptoms, stroke symptoms, and modifiable risk factors was low, i.e., less than 20% of the participants provided >75% correct responses to knowledge-related questions. Despite the low composite knowledge scores, most participants recognized chest pain and pain in arms or shoulder as common symptoms of heart attack. Many participants also identified numbness or weakness of the face, arm, or leg, or sudden confusion or trouble speaking or walking as symptoms of stroke. Further, this study showed that NPHW facilitated care and text messages for healthy lifestyle and risk factor monitoring to prevent or manage CVD are generally acceptable in a country with extremely limited specialist doctors and low physician-to-patient ratio.

When compared to people with existing CVD, lower level of knowledge of heart attack/stroke symptoms and risk factors was found predominantly in people with CVRFs, unskilled workers, low education, and low-income groups. Our study findings are consistent with previous reports. For example, a study of 444 adults from South Korea found the mean knowledge score of 4.3/9, 5.8/9, and 7.3/11, for heart attack symptoms, stroke symptoms, and modifiable risk factors, respectively^23^. Further, the regression analyses showed that older age and lower levels of education and income were associated with lower knowledge scores. In the current study, high blood cholesterol, high blood pressure, and stress/depression were identified as the most common modifiable risk factors for heart attack, which is consistent with studies from Nepal, Bangladesh, Pakistan, Iran, and other regions^24^. In this study, the knowledge of modifiable risk factors (>75% correct responses) was 17.3%, which is consistent with a study from Uganda involving 4,372 participants which found that a relatively small proportion of the respondents (18%) had good knowledge of CVD prevention. Although the survey instrument used was different, this study from Uganda revealed that education levels, occupation, household income, and social-economic index were significantly associated with the CVD knowledge^25^.

Several large epidemiological studies have shown a strong association between socio-economic position and CVD incidence and deaths. For instance, the Prospective Urban Rural Epidemiology (PURE) study analysed data of 160,299 adults aged 35-70 years old from urban and rural communities in 20 countries. After adjustment for wealth and other cofounding factors, the researchers found that low levels of education vs. high level of education was significantly associated with all-cause deaths: Hazard rate (HR), 95% CIs: 1.50 (95% CI: 1.14, 1.98) for high-income countries, 1.80 (95% CI: 1.58, 2.06) in middle-income countries, and 2.76 (95% CI: 2.29, 3.31) in low-income countries^26^. Further, O’Donnell et al. showed that sub-optimal knowledge, diagnosis, and treatment of hypertension were significantly associated with higher risk of stroke, younger age of stroke onset, and larger proportion of intracerebral haemorrhagic stroke in lower-income countries. This study underscored the potential role of population-wide strategies to improve knowledge of hypertension, and access to opportunistic screening for risk factors, to reduce the burden of premature stroke in LMICs^27^.

This study found that two-thirds of respondents were willing to receive NPHW delivered care for lifestyle modification advice related to healthy diet, exercise, and managing medicines as well as clinic visit reminders. Further, most participants were willing to receive text-messages for healthy lifestyle and to improve medication adherence, which is consistent with prior studies^28^. However, willingness to receive text-messages may not alone translate into adoption of healthy behaviours in long-term. Task-sharing and mHealth based strategies involving structured lifestyle modification programs to support patients have been extensively evaluated for its effectiveness in high-income countries to improve patient outcomes with some pilot studies from low- and middle-income countries^29^. A 2019 systematic review and meta-analysis of 31 studies found that task-sharing interventions with NPHW significantly reduced systolic and diastolic blood pressures (mean difference, 95% CI): -4.85 mm Hg (95% CI: -6.12 to -3.57), and -2.92 mm Hg (95% CI: - 3.75 to -2.09), respectively, in low- and middle-income countries^30^. Another study reported that a NPHW-led educational intervention improved adherence to guideline directed medical therapy and healthy lifestyle among patients with the acute coronary syndrome^31^. Park et al. reviewed 28 studies that applied mobile phone interventions (i.e., text messaging, mobile apps, tele-monitoring via mobile phones) for CVD management and found that text messaging appears to be more effective than smartphone interventions (e.g., mobile apps and telemonitoring) in secondary prevention of CVD^32^. Furthermore, a 2017 systematic review of 27 randomised trials found that mobile health interventions increased medication adherence, achievement of blood pressure targets, exercise, and increased awareness of diet and exercise for the secondary prevention of CVD^33^. A recent cross-sectional survey among 4,372 adults in Mukono and Buikwe districts in Uganda reported that participants who had received advice on healthy lifestyle through mobile phones had higher CVD knowledge as compared to those who had ever received healthy lifestyle advice; adjusted prevalence ratio = 1.37 (95% CI: 1.14, 1.65)^25^. However, data from recent trials of text-messaging showed null results^34,35^.

### Implications for clinical practice and policy

Given that increasingly CVD affects people in their economically most productive ages, these results have important implications for clinical practice and CVD prevention policies to minimize the adverse consequences of inadequate knowledge such as delayed diagnosis and treatment, increased disease burden due to uncontrolled risk factors, higher healthcare costs, reduced quality of life and social and economic challenges, all exacerbating existing health inequalities. Promoting awareness and adopting healthy behaviors to prevent and manage CVD involves a multi-faceted approach that targets individuals, communities, and healthcare systems. First, this study highlights the urgent need to implement community-based health education programs that provide information about CVD risk factors and preventive measures using various channels such as schools, workplaces, community centers, and social medial to reach out the target population at high risk of CVD. Second, to collaborate with media outlets to disseminate accurate and culturally appropriate information on CVD prevention, symptoms, and risk factors, as well as, engage in public service announcements to reach diverse populations. Third, train healthcare professionals to effectively communicate with patients about cardiovascular health, emphasizing the importance of early detection, risk factor control and lifestyle modifications during outpatient visits. Fourth, utilize technology to deliver health information and promote sustainable behavior change through mobile apps and interactive tools that provide personalized advice, track health metrics and offer resources for cardiovascular health. Lastly, policy advocacy that support cardiovascular health at the community and national levels through improved food labeling, urban planning that encourage physical activity, effective implementation of tobacco regulations, and regular health screenings to detect and manage CVD risk factors early.

### Strengths and limitations

This study has several strengths. First, we selected participants from a well-established, large cohort from Delhi and Chennai in North and South India using multi-stage cluster random sampling, and compared knowledge, attitude, and practices among well-defined groups, i.e., participants with prior CVD vs those with CVRFs. Second, a standardized pre-tested questionnaire was administered by trained study staff. Third, we also assessed attitudes and practices around CVD management to inform design of targeted prevention strategies, i.e., acceptability of NPHW delivered care and willingness to use text-messages, which are potential, yet under-studied strategies to improve knowledge-practice gaps in CVD prevention.

This study also has some limitations. This is an observational cross-sectional study that relied on self-reported data. The self-reported measures of modifiable risk factors may have overestimated the knowledge as the participants may have responded positively to the cardiovascular risk factors knowing that this study was focused on CVD (i.e., healthy volunteer bias). However, knowledge of heart attack and stroke symptoms related questions are unlikely to be affected. We did an exploratory analysis to determine the association of knowledge of CVD symptoms, and risk factors with socio-demographic factors. The reported confidence intervals for occupation category and low-income group, were wide, which may be due to small sample size. Also, data for risk factor levels (blood pressure, glucose, cholesterol, and weight) were obtained from 2016-17, which may not reflect participants’ current values. However, these data points were not included in the multivariable regression model and the study results remain unaffected by the use of old laboratory values. Participants were selected from two urban cities in India, which limits the generalizability of this study results to India’s other cities and its rural population. However, study participants had diverse representation across education and income groups. Although respondents’ health-seeking behaviours (self-monitoring of CVD risk factors) were relatively high, the risk of social desirability bias might have led the respondents to provide answers expected by interviewers. Further, given the study data collection period between January-February 2021, we cannot ascertain the influence of COVID-19 pandemic on the perceived knowledge, attitudes, and practices regarding CVD.

## Conclusion

This study shows substantial variation and sub-optimal knowledge of heart attack and stroke symptoms and CV risk factors in India. The use of task-sharing approaches, including trained NPHWs, and use of SMS-based reminders for healthy lifestyle were acceptable to most participants across major subgroups, which may improve the uptake of preventive CVD care. However, to engage and empower people at high CVD risk, culturally appropriate, and targeted educational programs leveraging digital technologies customized for elderly, low socio-economic groups would be desirable.

## Data Availability

Corresponding author has access to all study data. Data will be made available to the external researchers upon request.

## Disclosures (conflict of interest)

MDH has received travel support from the American Heart Association and World Heart Federation and consulting fees from PwC Switzerland. MDH has an appointment at The George Institute for Global Health, which has a patent, license, and has received investment funding with intent to commercialize fixed-dose combination therapy through its social enterprise business, George Medicines. MDH has pending patents for heart failure polypills.

---

**eFigure 1a.**
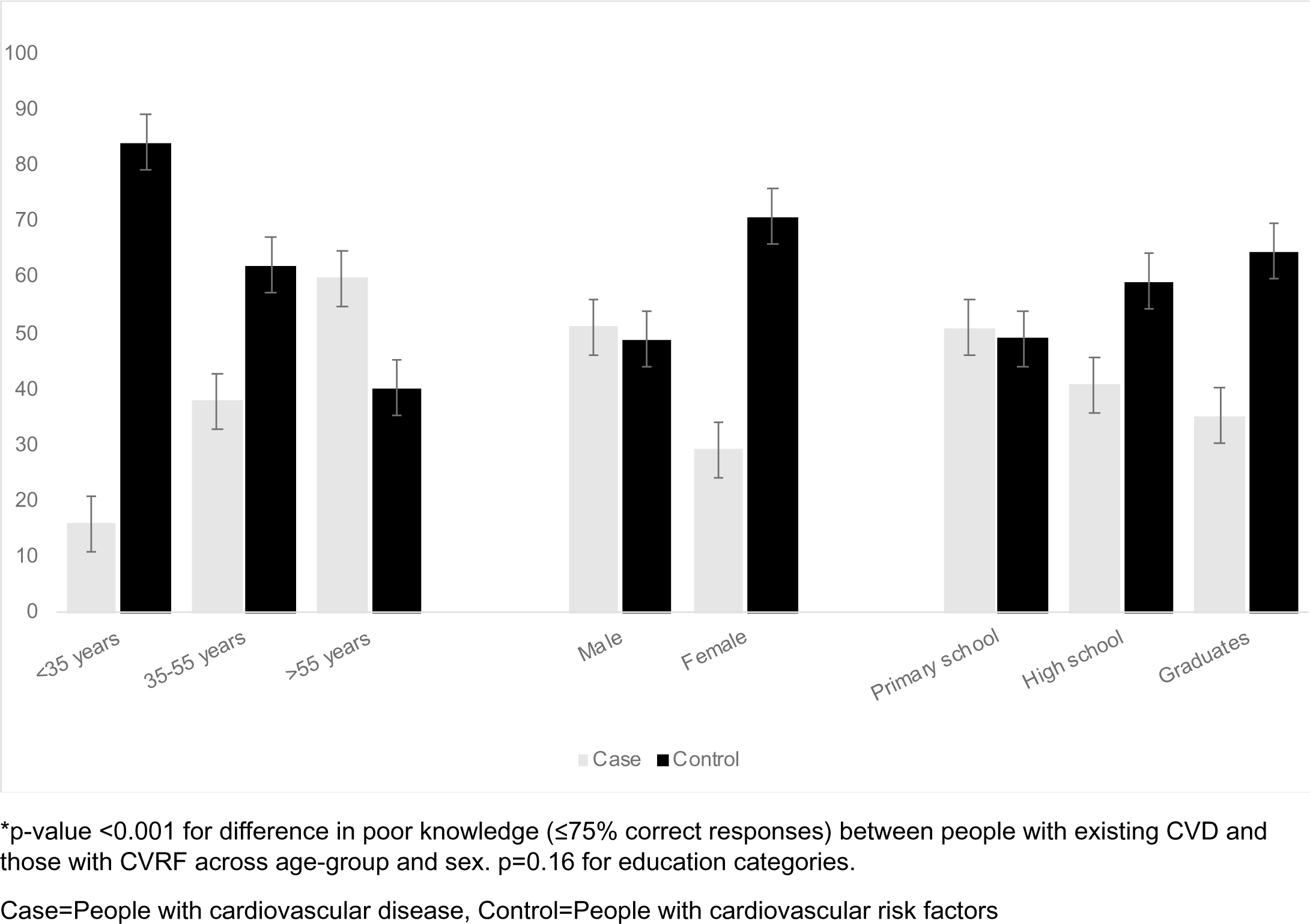
Lower level knowledge of heart attack symptoms among people with existing CVD vs people with CVRF by socio-demographic categories.

**eFigure 1b.**
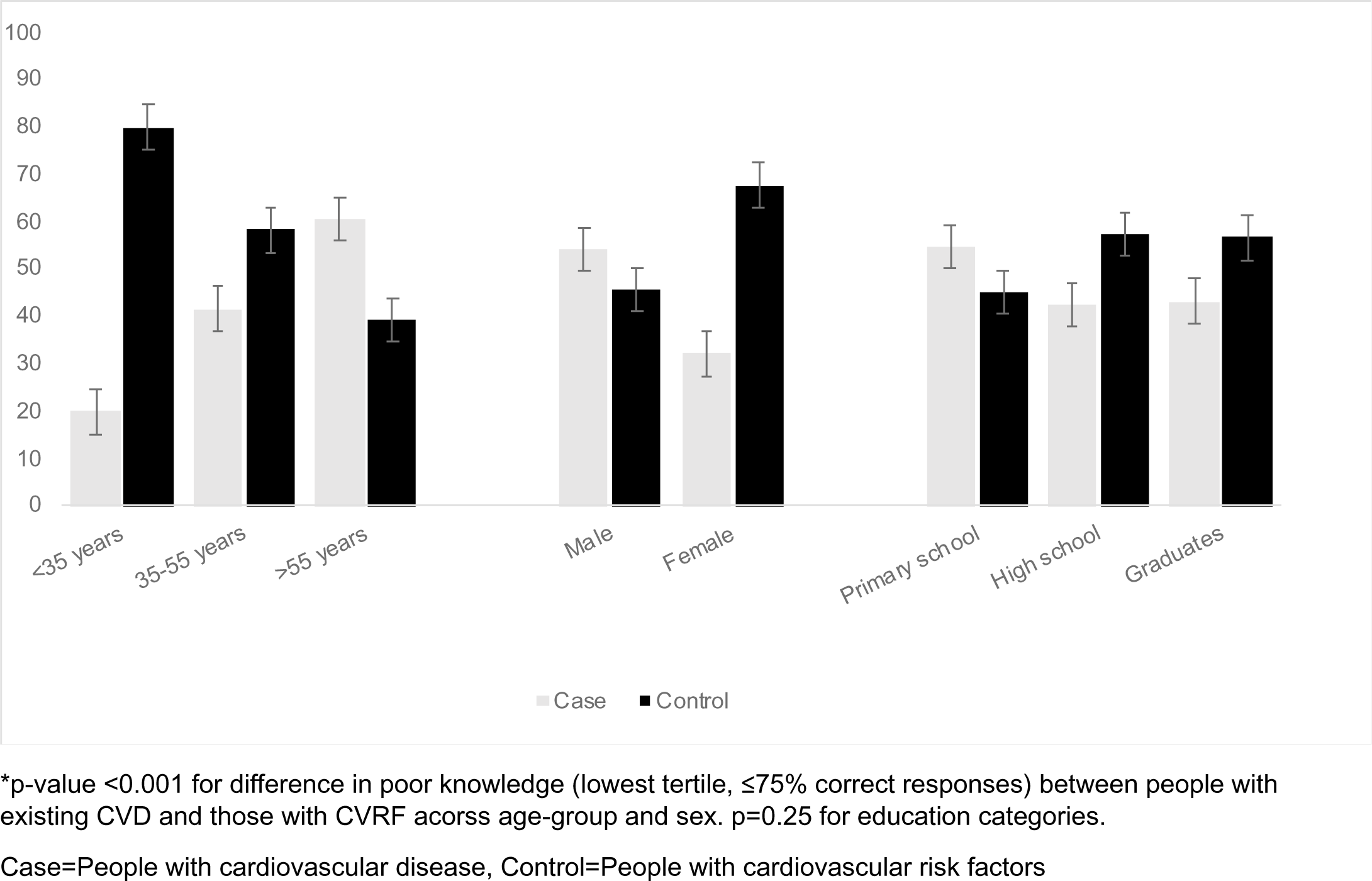
Lower level knowledge of stroke symptoms among people with existing CVD vs people with CVRF by socio-demographic categories.

**eFigure 1c.**
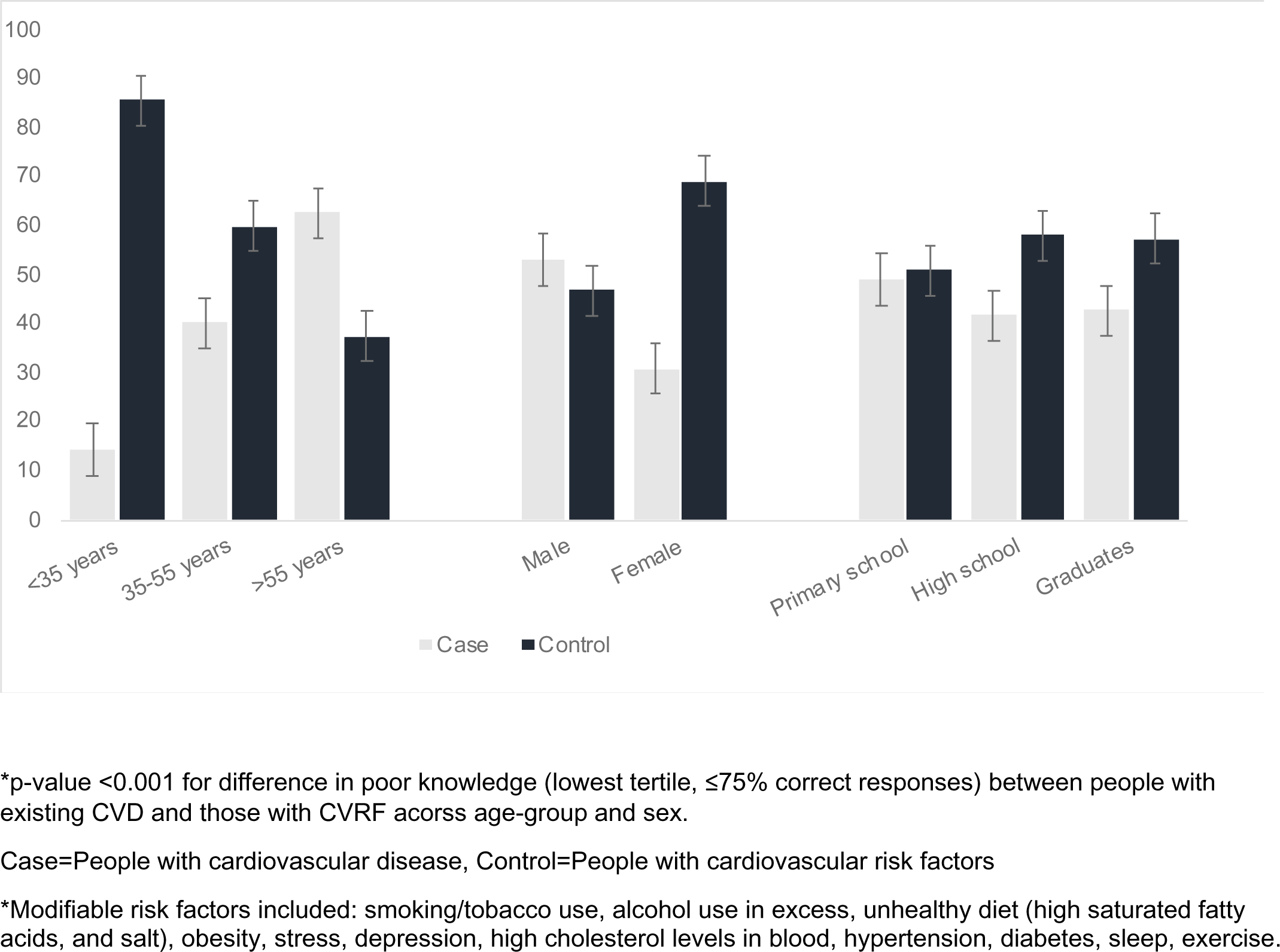
Lower level knowledge of modifiable risk factors among people with existing CVD vs people with CVRF by socio-demographic categories.

**eTable 1.** Practices related to monitoring of cardiovascular risk factors.

CVRF=Cardiovascular Risk Factors, CHD=coronary heart disease **eTable 1.** Practices related to monitoring of cardiovascular risk factors.
| Parameters | Overall<br>N=502 (%) | Control<br>N=283 (%) | Cases<br>N=219 (%) | P-value |
| --- | --- | --- | --- | --- |
| **Blood pressure checked last time** |  |  |  | <0.001 |
| Checked in the last 12 months | 374 (74.5%) | 192 (67.8%) | 182 (83.1%) |  |
| Checked >12 months ago | 108 (21.5%) | 76 (26.9%) | 32 (14.6%) |  |
| Never checked | 20 (4.0%) | 15 (5.3%) | 5 (2.3%) |  |
| **Blood cholesterol checked last time** |  |  |  | <0.001 |
| Checked in the last 12 months | 285 (56.8%) | 139 (49.1%) | 146 (66.7%) |  |
| Checked >12 months ago | 166 (33.1%) | 113 (39.9%) | 53 (24.2%) |  |
| Never checked | 51 (10.2%) | 31 (11.0%) | 20 (9.1%) |  |
| **Blood glucose checked last time** |  |  |  | <0.001 |
| Checked in the last 12 months | 348 (69.3%) | 176 (62.2%) | 172 (78.5%) |  |
| Checked >12 months ago | 124 (24.7%) | 89 (31.4%) | 35 (16.0%) |  |
| Never checked | 30 (6.0%) | 18 (6.4%) | 12 (5.5%) |  |
| **Body weight checked last time** |  |  |  | 0.007 |
| Checked in the last 12 months | 322 (64.1%) | 171 (60.4%) | 151 (68.9%) |  |
| Checked >12 months ago | 142 (28.3%) | 95 (33.6%) | 47 (21.5%) |  |
| Never checked | 38 (7.6%) | 17 (6.0%) | 21 (9.6%) |  |
| **Monitoring CVRF\*** |  |  |  | 0.083 |
| All four | 194 (38.6%) | 100 (35.3%) | 94 (42.9%) |  |
| Any one | 308 (61.4%) | 183 (64.7%) | 125 (57.1%) |  |
| **Recent blood pressure (perceived)** |  |  |  | <0.001 |
| Normal | 406 (80.9%) | 219 (77.4%) | 187 (85.4%) |  |
| High | 37 (7.4%) | 18 (6.4%) | 19 (8.7%) |  |
| Don't Know | 57 (11.4%) | 46 (16.3%) | 11 (5.0%) |  |
| **Recent blood cholesterol (perceived)** |  |  |  | <0.001 |
| Normal | 335 (66.7%) | 180 (63.6%) | 155 (70.8%) |  |
| High | 22 (4.4%) | 5 (1.8%) | 17 (7.8%) |  |
| Don't Know | 136 (27.1%) | 97 (34.3%) | 39 (17.8%) |  |
| **Recent fasting blood glucose(perceived)** |  |  |  | 0.019 |
| Normal | 369 (73.5%) | 202 (71.4%) | 167 (76.3%) |  |
| High | 49 (9.8%) | 24 (8.5%) | 25 (11.4%) |  |
| Don't Know | 81 (16.1%) | 57 (20.1%) | 24 (11.0%) |  |

**eTable 2.** Factors associated with low monitoring of CVD risk factors.

|  | Knowledge score |  | Unadjusted | Adjusted* |
| --- | --- | --- | --- | --- |
|  | High<br>N (%) | Low<br>N (%) | Odds ratio (95% CI) | Odds ratio (95% CI) |
| **Monitoring of CVD risk factors** |  |  |  |  |
| People with CVRF | 100 (35.3%) | 183 (64.7%) | 1.38 (0.96,1.98) | 1.50 (1.00,2.23) |
| Female | 89 (38.7%) | 141 (61.3%) | 1.00 (0.69,1.43) | 0.92 (0.63,1.34) |
| Age (in years) | 194 (38.7%) | 308 (61.3%) | 0.99 (0.97,1.00) | 0.99 (0.97,1.01) |
| Marital Status (Married) | 186 (39.4%) | 286 (60.6%) | 0.56 (0.24,1.28) | 0.57 (0.24,1.34) |
| Education |  |  |  |  |
| *Graduation and above* | 52 (43.7%) | 67 (56.3%) | ref | ref |
| *Up to primary schooling* | 20 (32.8%) | 41 (67.2%) | 1.59 (0.83,3.03) | 2.84 (1.39,5.78) |
| *High school to Secondary school* | 122 (37.9%) | 200 (62.1%) | 1.27 (0.83,1.95) | 2.07 (1.27,3.38) |
| Occupation |  |  |  |  |
| *Trained/skilled* | 70 (39.8%) | 106 (60.2%) | ref | ref |
| *Unemployed* | 95 (39.9%) | 143 (60.1%) | 0.99 (0.67,1.48) | 1.34 (0.76,2.36) |
| *Semiskilled/Unskilled* | 29 (33.0%) | 59 (67.0%) | 1.34 (0.79,2.30) | 1.77 (1.00,3.13) |
| Monthly household income (in Indian rupees) |  |  |  |  |
| >20,000 | 51 (40.5%) | 75 (59.5%) | ref | ref |
| <10,000 | 100 (36.8%) | 172 (63.2%) | 1.17 (0.76,1.80) | 2.02 (1.19,3.41) |
| 10,000-20,000 | 43 (42.2%) | 59 (57.8%) | 0.93 (0.55,1.59) | 1.25 (0.71,2.20) |
| Family history of CHD (No) | 163 (38.4%) | 261 (61.6%) | 1.06 (0.64,1.73) | 1.19 (0.71,1.99) |

\*The regression model is adjusted for city, age and sex. #monitoring of CVD risk factors includes blood pressure, blood glucose, and blood cholesterol
CVRF=Cardiovascular Risk Factors

## Notes

### Competing Interest Statement

Mark D Huffman has received travel support from the American Heart Association and World Heart Federation and consulting fees from PwC Switzerland. MDH has an appointment at The George Institute for Global Health, which has a patent, license, and has received investment funding with intent to commercialize fixed-dose combination therapy through its social enterprise business, George Medicines. MDH has pending patents for heart failure polypills.

### Clinical Trial

Not applicable

### Funding Statement

The CARRS cohort was funded by the National Heart Lung Blood Institute, National Institutes of Health, USA. Dr Kavita Singh is supported by the Fogarty International Centre, National Institutes of Health (NIH), United States (grant award: 1K43TW011164). The content is solely the responsibility of the authors and does not necessarily represent the official views of the National Institutes of Health.

### Author Declarations

Institutional Ethics Committees of Public Health Foundation of India and Madras Diabetes Research Foundation, Chennai, India.

## References

1. Roth GA, Mensah GA, Johnson CO, et al. Global Burden of Cardiovascular Diseases and Risk Factors, 1990-2019: Update From the GBD 2019 Study. J Am Coll Cardiol. Dec 22 2020;76(25):2982–3021. doi:10.1016/j.jacc.2020.11.010

2. Karan A, Engelgau M, Mahal A. The household-level economic burden of heart disease in India. Trop Med Int Health. May 2014;19(5):581–91. doi:10.1111/tmi.12281

3. Mondal S, Van Belle S. India’s NCD strategy in the SDG era: are there early signs of a paradigm shift? Global Health. Apr 25 2018;14(1):39. doi:10.1186/s12992-018-0357-6

4. Global Non-Communicable Disease (NCD) Action Plan. Accessed 10 Dec 2023. https://ncdalliance.org/global-ncd-action-plan.

5. World Health Organization (WHO) Global Action Plan 2019. Accessed on 10 Dec 2023. https://www.who.int/initiatives/sdg3-global-action-plan.

6. Vaidya A, Aryal UR, Krettek A. Cardiovascular health knowledge, attitude and practice/behaviour in an urbanising community of Nepal: a population-based cross-sectional study from Jhaukhel-Duwakot Health Demographic Surveillance Site. BMJ Open. Oct 24 2013;3(10):e002976. doi:10.1136/bmjopen-2013-002976

7. Amarasekara P, de Silva A, Swarnamali H, Senarath U, Katulanda P. Knowledge, Attitudes, and Practices on Lifestyle and Cardiovascular Risk Factors Among Metabolic Syndrome Patients in an Urban Tertiary Care Institute in Sri Lanka. Asia Pac J Public Health. Jan 2016;28(1 Suppl):32S–40S. doi:10.1177/1010539515612123

8. Khan MS, Jafary FH, Jafar TH, et al. Knowledge of modifiable risk factors of heart disease among patients with acute myocardial infarction in Karachi, Pakistan: a cross sectional study. BMC Cardiovasc Disord. Apr 27 2006;6:18. doi:10.1186/1471-2261-6-18

9. Islam FM, Chakrabarti R, Islam MT, et al. Prediabetes, diagnosed and undiagnosed diabetes, their risk factors and association with knowledge of diabetes in rural Bangladesh: The Bangladesh Population-based Diabetes and Eye Study. J Diabetes. Mar 2016;8(2):260–8. doi:10.1111/1753-0407.12294

10. Labarthe D, Lloyd-Jones DM. 50×50×50: Cardiovascular Health and the Cardiovascular Disease Endgame. Circulation. Sep 4 2018;138(10):968–970. doi:10.1161/CIRCULATIONAHA.118.035985

11. Anand SS, Samaan Z, Middleton C, et al. A Digital Health Intervention to Lower Cardiovascular Risk: A Randomized Clinical Trial. JAMA Cardiol. Aug 1 2016;1(5):601–6. doi:10.1001/jamacardio.2016.1035

12. Liu G, Li Y, Hu Y, et al. Influence of Lifestyle on Incident Cardiovascular Disease and Mortality in Patients With Diabetes Mellitus. J Am Coll Cardiol. Jun 26 2018;71(25):2867–2876. doi:10.1016/j.jacc.2018.04.027

13. Palmer MJ, Barnard S, Perel P, Free C. Mobile phone-based interventions for improving adherence to medication prescribed for the primary prevention of cardiovascular disease in adults. Cochrane Database Syst Rev. Jun 22 2018;6:CD012675. doi:10.1002/14651858.CD012675.pub2

14. Ali MK, Singh K, Kondal D, et al. Effectiveness of a Multicomponent Quality Improvement Strategy to Improve Achievement of Diabetes Care Goals: A Randomized, Controlled Trial. Ann Intern Med. Sep 20 2016;165(6):399–408. doi:10.7326/M15-2807

15. Nair M, Ali MK, Ajay VS, et al. CARRS Surveillance study: design and methods to assess burdens from multiple perspectives. BMC Public Health. Aug 28 2012;12:701. doi:10.1186/1471-2458-12-701

16. Kondal D, Patel SA, Ali MK, et al. Cohort Profile: The Center for cArdiometabolic Risk Reduction in South Asia (CARRS). Int J Epidemiol. Feb 9 2022;doi:10.1093/ije/dyac014

17. Chobanian AV, Bakris GL, Black HR, et al. The Seventh Report of the Joint National Committee on Prevention, Detection, Evaluation, and Treatment of High Blood Pressure: the JNC 7 report. JAMA. May 21 2003;289(19):2560–72. doi:10.1001/jama.289.19.2560

18. Use of Glycated Haemoglobin (HbA1c) in the Diagnosis of Diabetes Mellitus: Abbreviated Report of a WHO Consultation. 2011. WHO Guidelines Approved by the Guidelines Review Committee.

19. Genuth S, Alberti KG, Bennett P, et al. Follow-up report on the diagnosis of diabetes mellitus. Diabetes Care. Nov 2003;26(11):3160–7. doi:10.2337/diacare.26.11.3160

20. Saeed O, Gupta V, Dhawan N, et al. Knowledge of modifiable risk factors of Coronary Atherosclerotic Heart Disease (CASHD) among a sample in India. BMC Int Health Hum Rights. Feb 4 2009;9:2. doi:10.1186/1472-698X-9-2

21. Indian Council of Medical Research (ICMR) guidelines on health research during COVID-19 Pandemic in India. Accessed on 20 Mar 2022. https://www.icmr.gov.in/pdf/covid/techdoc/EC_Guidance_COVID19_06052020.pdf.

22. Commcare Data Collection App. Accessed on 20 March 2022. https://www.dimagi.com/commcare/.

23. Han CH, Kim H, Lee S, Chung JH. Knowledge and Poor Understanding Factors of Stroke and Heart Attack Symptoms. Int J Environ Res Public Health. Sep 29 2019;16(19)doi:10.3390/ijerph16193665

24. Koohi F, Khalili D. Knowledge, Attitude, and Practice Regarding Cardiovascular Diseases in Adults Attending Health Care Centers in Tehran, Iran. Int J Endocrinol Metab. Jul 2020;18(3):e101612. doi:10.5812/ijem.101612

25. Ndejjo R, Nuwaha F, Bastiaens H, Wanyenze RK, Musinguzi G. Cardiovascular disease prevention knowledge and associated factors among adults in Mukono and Buikwe districts in Uganda. BMC Public Health. Jul 22 2020;20(1):1151. doi:10.1186/s12889-020-09264-6

26. Rosengren A, Smyth A, Rangarajan S, et al. Socioeconomic status and risk of cardiovascular disease in 20 low-income, middle-income, and high-income countries: the Prospective Urban Rural Epidemiologic (PURE) study. Lancet Glob Health. Jun 2019;7(6):e748–e760. doi:10.1016/S2214-109X(19)30045-2

27. M OD, Hankey GJ, Rangarajan S, et al. Variations in knowledge, awareness and treatment of hypertension and stroke risk by country income level. Heart. Dec 14 2020;doi:10.1136/heartjnl-2019-316515

28. DeSouza SI, Rashmi MR, Vasanthi AP, Joseph SM, Rodrigues R. Mobile phones: the next step towards healthcare delivery in rural India? PLoS One. 2014;9(8):e104895. doi:10.1371/journal.pone.0104895

29. Singh K, Bawa VS, Venkateshmurthy NS, et al. Assessment of Studies of Quality Improvement Strategies to Enhance Outcomes in Patients With Cardiovascular Disease. JAMA Netw Open. Jun 1 2021;4(6):e2113375. doi:10.1001/jamanetworkopen.2021.13375

30. Anand TN, Joseph LM, Geetha AV, Prabhakaran D, Jeemon P. Task sharing with non-physician health-care workers for management of blood pressure in low-income and middle-income countries: a systematic review and meta-analysis. Lancet Glob Health. Jun 2019;7(6):e761–e771. doi:10.1016/S2214-109X(19)30077-4

31. Sharma KK, Gupta R, Mathur M, et al. Non-physician health workers for improving adherence to medications and healthy lifestyle following acute coronary syndrome: 24-month follow-up study. Indian Heart J. Nov - Dec 2016;68(6):832–840. doi:10.1016/j.ihj.2016.03.027

32. Park LG, Beatty A, Stafford Z, Whooley MA. Mobile Phone Interventions for the Secondary Prevention of Cardiovascular Disease. Prog Cardiovasc Dis. May-Jun 2016;58(6):639–50. doi:10.1016/j.pcad.2016.03.002

33. Gandhi S, Chen S, Hong L, et al. Effect of Mobile Health Interventions on the Secondary Prevention of Cardiovascular Disease: Systematic Review and Meta-analysis. Can J Cardiol. Feb 2017;33(2):219–231. doi:10.1016/j.cjca.2016.08.017

34. Klimis H, Thiagalingam A, McIntyre D, Marschner S, Von Huben A, Chow CK. Text messages for primary prevention of cardiovascular disease: The TextMe2 randomized clinical trial. Am Heart J. Dec 2021;242:33–44. doi:10.1016/j.ahj.2021.08.009

35. Chow CK, Klimis H, Thiagalingam A, et al. Text Messages to Improve Medication Adherence and Secondary Prevention After Acute Coronary Syndrome: The TEXTMEDS Randomized Clinical Trial. Circulation. May 10 2022;145(19):1443–1455. doi:10.1161/CIRCULATIONAHA.121.056161

